# Clustering fibromyalgia patients: A combination of psychosocial and somatic factors leads to resilient coping in a subgroup of fibromyalgia patients

**DOI:** 10.1101/2020.07.08.20148130

**Authors:** Alexandra Braun, Dimitar Evdokimov, Johanna Frank, Paul Pauli, Nurcan Üçeyler, Claudia Sommer

## Abstract

**Background:** Coping strategies and their efficacy vary greatly in patients suffering from fibromyalgia syndrome (FMS).

**Objective:** We aimed to identify somatic and psychosocial factors that might contribute to different coping strategies and resilience levels in FMS.

**Subjects and methods:** Standardized questionnaires were used to assess coping, pain, and psychological variables in a cohort of 156 FMS patients. Quantitative real-time polymerase chain reaction (qRT-PCR) determined gene expression of selected cytokines in white blood cells of 136 FMS patients and 25 healthy controls. Data of skin innervation, functional and structural sensory profiles of peripheral nociceptive nerve fibers of a previous study were included into the statistics. An exploratory factor analysis was used to define variance explaining factors, which were then included into cluster analysis.

**Results:** 54.9% of the variance was explained by four factors which we termed (1) affective load, (2) coping, (3) pain, and (4) pro-inflammatory cytokines (p < 0.05). Considering differences in the emerged factors, coping strategies, cytokine profiles, and disability levels, 118 FMS patients could be categorized into four clusters which we named “maladaptive”, “adaptive”, “vulnerable”, and “resilient” (p < 0.05). The adaptive cluster had low scores in disability and in all symptom categories in contrast to the vulnerable cluster, which was characterized by high scores in catastrophizing and disability (p < 0.05). The resilient vs. the maladaptive cluster was characterized by better coping and a less pro-inflammatory cytokine pattern (p < 0.05).

**Conclusion:** Our data suggest that problem- and emotion-focused coping strategies and an anti-inflammatory cytokine pattern are associated with reduced disability and might promote resilience. Additional personal factors such as low anxiety scores, ability of acceptance, and persistence further favor a resilient phenotype. Individualized therapy should take these factors into account.

## Introduction

Fibromyalgia syndrome (FMS) is a chronic pain syndrome with specific associated symptoms [1]. FMS patients show heterogeneity in symptoms and in coping with the disease’s impact on daily activities and professional life [2-7]. Some patients deal actively and effectively with pain and accompanying FMS symptoms, while others are vulnerable and severely disabled [8]. Activating resources and using effective coping with the disease might also be named resilience. The term resilience stems from the Latin word “*resilire*” meaning “bouncing back” from difficult experiences and is related to the phenomenon of psychological resistance despite intense distress and high risks [9, 10]. One definition of resilience is a flexible ability to adapt to every situation in the best fitting biopsychosocial way [11, 12]. Other definitions understand resilience as a personal trait or characteristic [13] that allows to activate resources. Better knowledge of the factors involved in resilience might guide individualized therapies.

Psychosocial factors that are well known to contribute to a resilient profile are among others solution-focused and active coping strategies and supportive social contacts [14]. Resilient people show low scores in depression, anxiety, and pain catastrophizing and remain with low disability after traumatic events and stressful life periods [15]. Decades of research in the field of resilience found biological determinants, which allowed to identify neural circuits and signaling pathways that mediate resilient phenotypes [16, 17]. Previous studies from our group showed that patients with widespread chronic pain have lower levels of the anti-inflammatory cytokines interleukin (IL)-4 and IL-10 compared to healthy controls [18]. Accordingly, in patients with polyneuropathies, higher blood levels of IL-4 and IL-10 were associated with a painless phenotype [19]. Recently, we showed that a subgroup of FMS patients with markedly reduced skin innervation, i.e. with pathological peripheral nociceptors, has more severe FMS symptoms [20, 21]. Here we wanted to assess, whether psychosocial factors, the cytokine profile, or the state of peripheral nociceptive nerve fibers as measured by functional and structural tests, are associated with successful coping and resilience in FMS.

Given the wide range of variables, we used an exploratory factor analysis of this clinically well-defined FMS patient cohort with the aim to find independent factors explaining the high variance within the cohort. Given the results from our previous studies, we included gene expression data of pro- and anti-inflammatory cytokines, physiological data of quantitative sensory testing (QST) and pain-related evoked potential (PREP) measurements, and data of intraepidermal nerve fiber density (IEFND) derived from skin punch biopsies as somatic variables. Psychosocial data were derived from the medical history and a series of standardized questionnaires evaluating depression, anxiety, pain catastrophizing, early life stress, and the way of coping. The following hierarchical clustering aimed to identify subgroups with differences in these predefined factors and characterize them regarding biopsychosocial aspects, severity levels, and resilient or vulnerable phenotypes, with the goal to identify resilient coping patterns. In the long run, these patterns might be used to develop individualized therapies for the subgroups.

Several authors have performed cluster analyses to identify subgroups among fibromyalgia patients characterized by symptom severity, objective or subjective measures of physical and mental health with different outcomes [22-27]. One study stands out with a large cohort of 947 FMS patients assessed with the FIQ-R sum score variable, resulting in four clusters with differences in clinical, economical and somatic outcomes like inflammatory markers and grey matter volume [28]. In contrast to this study [28], which did clustering only with a single sum score variable, our approach was to cluster the cohort by previously emerged validated factors, a method that is supposed to make data more transferrable on other cohorts independently on regional differences.

## Materials and Methods

### Subjects and ethics

This study is part of a current larger study on FMS at the Department of Neurology of the University Hospital Würzburg, in course of which 156 FMS patients fulfilling the diagnostic criteria for FMS of the American College of Rheumatology published in 2010 [29], and 48 age and gender matched healthy controls were recruited between 2014 to 2018 [20]. Exclusion and inclusion criteria were as specified elsewhere [30] (see S1 Table). All patients provided written informed consent and filled in a series of questionnaires before enrollment. An overview of the examination plan of the main study is included (see S2 Fig). Data on clinical examination, electrophysiological and laboratory measurements, and the tests in the ophthalmology department were published elsewhere [20, 21]. Details among clusters are summed up in S3 Table. Our study was approved by the Würzburg University Ethics Committee (No. 135/15).

### Psychosocial and symptom specific questionnaires as variables for clustering

#### Psychosocial variables

All questionnaires were available as standardized validated German versions as indicated by an added “D” at the end of the questionnaire abbreviation.

The Center of Epidemiological Studies General Depression Scale (CES-D) is a 20-items questionnaire that examines the severity of depressive symptoms. A total score of CES-D ≥ 22 is supposed to indicate clinically relevant depressive symptoms [31]. The State-Trait Anxiety Inventory (STAI-G) examines anxiety on a 1 to 4 scale as a trait (STAI-T) and as a state (STAI-S) shown by two sum scores [32]. The STAI-S sum score is created by the scores of item 1 to 20, and the STAI-T sum score by the items 21 to 40. The Pain Catastrophizing Scale (PCS-D) examines the strength of catastrophizing feeling and behavior [33]. The PCS sum score was created with the sum of all 13 items. The short form of the Childhood Trauma Questionnaire (CTQ-D) is a 28-items questionnaire that examines the experience (as frequency) and severity of traumatic events summarized in the five sub scales “sexual abuse”, “emotional abuse”, “emotional neglect”, “physical abuse”, physical neglect, and an additional scale of trivialization. The sum score of every subscale is 5 to 25 which is indicated as “severe” traumatic event [34, 35].

#### Coping

We used the Coping Strategies Questionnaire (CSQ-D) and its 8 subscales (distraction of attention, reinterpretation, self-instructions, ignoring, praying and hoping, catastrophizing, increase in activity, pain behavior) plus the effectiveness ratings on pain control and pain reduction) to examine different strategies to cope with pain. The maximum sum score was 36 for each coping strategy and 6 for the effectiveness ratings [36].

#### Pain and FMS related disability

Three questionnaires assessed different aspects of pain and disability due to pain. The Neuropathic Pain Symptom Inventory (NPSI-D) examines the severity and characteristics of pain in the last 24 hours, which results in the NPSI sum score [37].The Graded Chronic Pain Scale (GCPS-D) is a 7-item questionnaire that calculates 3 sub scores showing pain intensity, disability due to pain and the grade of disability, which is constructed from a combination of a disability scale from 0 to over 70 and days of disability from 0 to 31 days [38].The Fibromyalgia Impact Questionnaire (FIQ-D) examines physical functioning, working status, well-being of FMS patients, and their impact on daily life [39]. The Short-Form Health Survey (SF-12) is the shorter version of the SF-36 questionnaire and a screening tool for measuring health-related quality of life [40, 41]. The Physical Component Summary (PCS) scale represents general health perception, physical function and role function as well as pain. The Mental Component Summary (MCS) scale represents emotional role function, mental well-being, negative affect and social functioning. The FIQ sum score and both subscales of the SF-12 represent physical disability and quality of life and was used as component to define the severity level of affection resulting in low or high resilience in our cohort.

### Somatic data as variables for clustering

#### Blood withdrawal, white blood cell extraction, and RNA isolation

Venous blood was collected from all patients and 25 healthy controls between 8:00 AM and 9:00 AM after overnight fasting. Subjects were instructed not to consume alcohol, not to smoke and not to perform strenuous physical activity 24 h before the study day. Two serum monovettes (4.7 ml) and three ethylendiamine-tetra-acetic acid-containing tubes (EDTA, 9.0 ml; Sarstedt AG & Co., Nümbrecht, Germany) were taken. After incubation for 30 minutes on ice, the white blood cell (WBC) fraction was extracted and 9 falcon tubes were filled with 1.5 ml EDTA blood each and 7.5 ml erythrocyte lysis buffer (EL buffer; Qiagen, Hilden, Germany) was added and gently mixed. Afterwards the second incubation on ice for 30 minutes was interrupted for 3 times vortexing. All 9 falcon tubes were centrifuged (400 g) for 10 minutes at +4°C, supernatant was discarded, and each cell pellet resuspended in 3 ml EL buffer. The same centrifugation step was conducted, and the supernatant discarded. The total white blood cell (WBC) fraction of 8 falcon tubes was resuspended in RNA Protect Cell Reagent (Qiagen, Hilden, Germany) and stored at −80°C until PCR analyses. 1 ml EL buffer was added to the 9^th^ falcon tube and 1 µl of the content was added to a Neubauer improved cell chamber (Bürker, 0.100 mm depth, 0.0025 mm^2^; BLAUBRAND®, Brand GmbH & Co.KG, Wertheim, Germany), and cells were quantified. After extraction, plasma and serum samples were frozen at −80°C, and the mRNA was isolated from 136 WBC samples following the manufacturer’s protocol using the miRNeasy Mini kit (Qiagen, Hilden, Germany).

#### cDNA synthesis

For reverse transcription of 250 ng of the isolated mRNA to cDNA, TaqMan Reverse Transcription Reagents^®^ (Applied Biosystems, Darmstadt, Germany) were used as previously described [87]. The cycler (ABI PRISM 7700 Cycler, Applied Biosystems, Darmstadt, Germany) was set as annealing at 25°C for 10 minutes, reverse transcription at 48°C for 60 minutes and enzyme inactivation at 95°C for 5 minutes.

#### TaqMan qRT − PCR

5 μl of transcribed cDNA was used for qRT-PCR performed in GeneAmp 7700 sequence detection system^®^ (Applied Biosystem, Darmstadt, Germany) as previously described [19]. Gene specific TaqMan primers for IL-6 (assay-ID: Hs00174131_m1), tumor necrosis factor (TNF) (assay-ID: Hs00174128_m1), IL-4 (assay – ID: Hs00174122_m1), IL-10 (assay-ID: Hs00174086_m1), and endogenous control 18sRNA as well as a WBC control from human blood of a healthy person were used for the TaqMan Gene Expression Assays^®^ (Applied Biosystems, Darmstadt, Germany). Samples on each PCR plate were run as triplicates, endogenous control samples were run as duplicates. Each plate contained a negative control (RNA free water) to exclude genomic contamination. Before the samples run, control samples were used to verify a calibrator as a standard sample on each plate to guarantee inter-plate comparability. For evaluation of relative gene expression of all four target genes compared between patients and controls we used the comparative deltaCT method (lower deltaCT values express higher gene expression). As a more convenient visualization of the results we calculated 1/deltaCT to illustrate higher values as higher gene expression (compare [42]).

#### Further physiological data

Data of quantitative sensory testing (QST), pain-related evoked potential (PREP) measurements, data from clinical examination and history, and data of intraepidermal nerve fiber density (IENFD) derived from skin biopsies of upper and lower leg were collected in the main study and here used as variables for multivariate variance analyses (factoring and clustering). These data were published elsewhere [20, 21] and summarized in S3 Table regarding patient cluster of the current study (see S3 Table).

#### Statistical analysis

IBM SPSS Statistics 25 software (IBM, Ehningen, Germany) was used for statistical analysis. Data distribution was tested with the Shapiro-Wilk test and by observing data histograms. Results of the non-normally distributed data are given as median (MED) and range, and of normally distributed data as mean (M) and standard deviation (SD).

##### Z transformation

Sum scores of the questionnaires and somatic data used in factor analysis were saved as standardized values to make all variables comparable independent of scaling in the total data sheet within SPSS.

##### Higher order principal factor analysis (PFA)

Extraction method was a principal axis factoring with oblimin rotation. The Kaiser–Meyer–Olkin (KMO = 0.77) measure verified sampling adequacy for the analysis and the Bartlett’s test of sphericity (χ^2^ (253) = 1322.276, p < 0.001) indicated that correlations between items were sufficiently large and not multicollinear for PFA (see S5 Table). The determinant of the R correlation matrix was higher than 0.00001, which implies no severe multicollinearity [43]. An initial analysis was run to obtain eigenvalues for each factor. Seven factors had eigenvalues of at least one verified by Kaiser’s criterion and in combination explained 70% of the variance (see S6 Table). The scree plot was slightly ambiguous and showed less inflexions starting at factor 3 to 7 (see S7A Fig). Given the large variable size of 23, and the convergence of the scree plot and Kaiser’s criterion on four factors, this is the number of factors that were retained in the final analysis (overall Cronbach’ alpha = 0.7, which is suitable for items regarding ability tests [44]). Variable loadings of < 0.30 were suppressed in the PFA. Factor scores were created by regression within the SPSS software. Internal consistency of factors was analyzed by Cronbach’s α. Missing cases were excluded listwise.

##### Cluster analysis

Using factor scores a hierarchical cluster analysis with Ward’s method was conducted to group the entire cohort to clusters specified by previous defined factorial characteristics. Visual inspection of the dendrogram (see S8 Fig) was done to indicate the number of clusters that should be considered. The decision on the number of clusters was additionally made following Rehm et al. [45] and van Leeuwen et al. [46] by practical considerations (i.e. the least frequent cluster should include a minimum of 15% of the total sample) and by heuristic interpretability of mean factor scores within clusters (i.e. agglomeration schedule within SPSS software). By using this combination of interpretability, the developed cluster solution minimizes the within-group variability and maximizes the between-group differences [47, 48]. With a defined number of four clusters, the hierarchical method was run again with fixed and saved cluster solution (range 2 to 5 cluster). The 3D illustration model of these cluster and their differences in the emerged factors were created by MATLAB (*MATrix LABoratory*, The MathWorks Inc., Natick, Massachusetts, USA) [49].

##### Differences between patient profiles

A one-way analysis of variance (ANOVA) and Games-Howell post-hoc analyses of the pairwise comparisons of the ANOVA were used to detect between-cluster differences in all variables included into the multivariate analyses.

##### Grades of severity

For symptom severity and evaluation of subgroups, we calculated effect sizes using Cohen’s d [50] to choose the resilient and the vulnerable profiles (using the mean and standard deviation of these two subgroups). The calculated effect sizes were interpreted as small (0.2), medium (0.5) or large (0.8) [51]. Level of significance was set at p < 0.05.

## Results

### Data availability

Questionnaire data were available for 136 patients except for the CTQ-D and SF-12-D, which was only available from 112 patients. Some questionnaires were added at a later stage, therefore only data of 118 patients were valid for the multivariate analyses (see S4 Fig). Controls were only included to validate the PCR data, the other parts of the study focuses on differences among FMS patients. Sufficient data of 136 patients and 25 healthy controls were available to include them into the PCR study. Overall, fewer patients and controls could be included in the PCR analysis because not all patients had blood samples available.

### Variance within psychosocial data, coping strategies, pain variables, and immune agents

Psychosocial data (CES-D, STAI-G-D, PCS-D, CTQ-D), coping strategies (CSQ-D), pain variables (GCPS-D, NPSI-D), skin innervation, and relative cytokine gene expression of all gene targets showed a large variability. As an example, the relative gene expression of all analyzed cytokines showed no intergroup difference between FMS patients and controls but had high within group variabilities (Fig 1).

**Fig 1:**
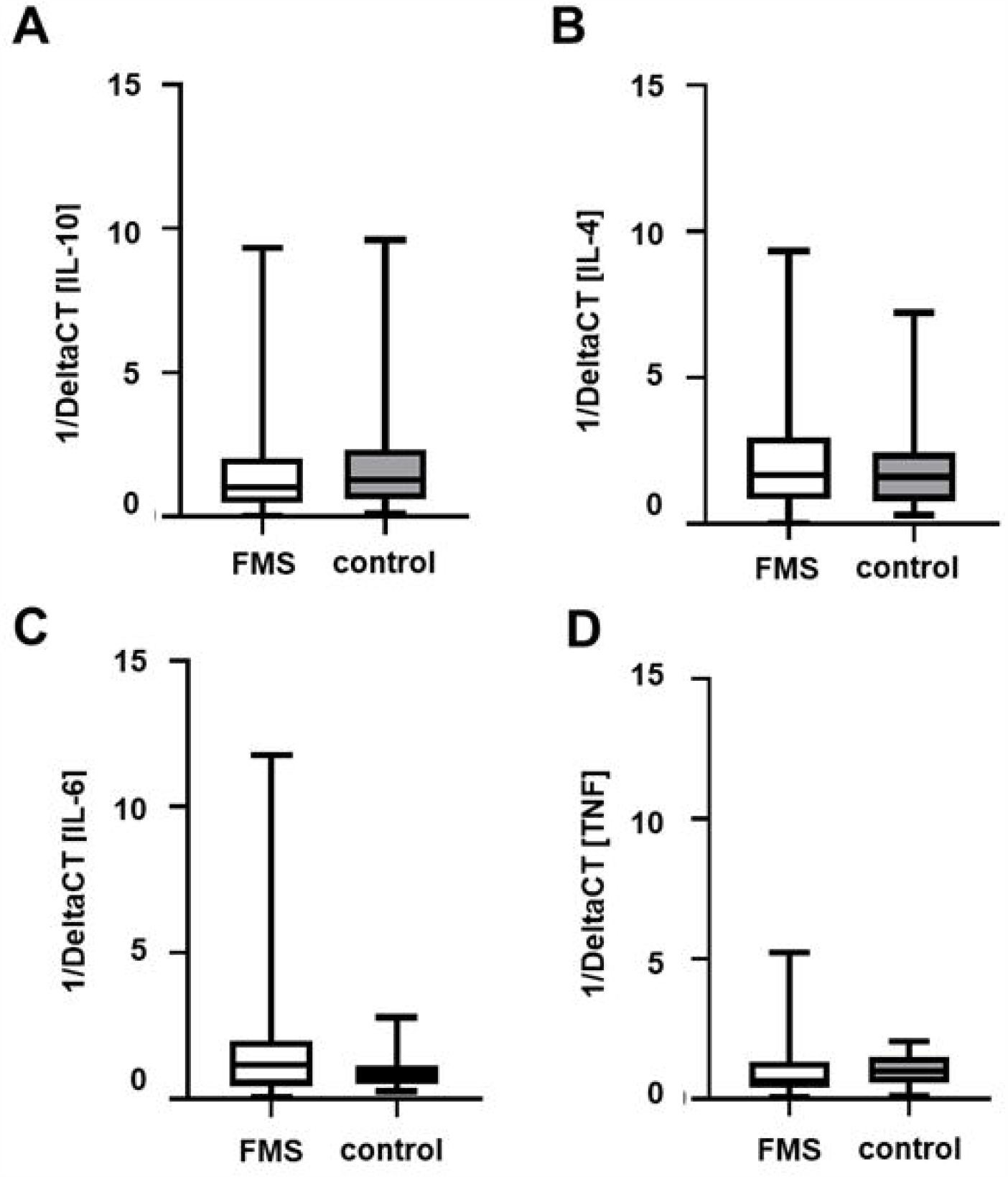
Relative gene expression of selected anti- and pro-inflammatory cytokines in FMS patients and healthy controls. Boxplots show deltaCT values, i.e. relation of the CT value of the target normalized to the housekeeping gene 18sRNA, of IL-4 (A), IL-6 (B), IL-10 (C) and TNF (D) of patients with FMS and healthy controls. Data are presented as 1/ΔCT. No intergroup difference was found for any of the investigated targets. Note the high variability within the patient group. Abbreviations: CT = cycle threshold; FMS = Fibromyalgia syndrome; IL = Interleukine; TNF = Tumor necrosis factor-alpha.

This overall huge variance and the clinical experience with this FMS patient cohort led us to assume the existence of more than two subgroups within the entire patient cohort. Due to the differences in a high amount of variables we aimed to exploratively test a range of variables to group the cohort.

### Mental health, coping, symptom disability and cytokine profile is responsible for variance

First, a factor analysis was performed on the data of 118/156 (75.6%) patients valid for multivariate analysis (see S5 Fig).

An initial run of the PFA was applied on 53 z -standardized variables to reduce the number of variables and to search for factors that explain the variance within the data field. 29 variables were identified as not responsible for the variance and not sufficient to be included in the final analysis. Factors related to nociceptor structure and function like proximal and distal IEFND did not emerge in the factor analysis. Data of quantitative sensory testing (QST) and evoked potentials nonspecifically loaded on two factors and could not be interpreted resulting in the decision to exclude them. After exclusion of these 29 variables, we ran a second PFA with 23 variables. The decision about the number of factors was based on inspection of the scree plot (see S7B Fig). Table 1 shows the factor loadings after rotation.

**Table 1:** Pattern matrix with factor loadings of every variable of the principal factor analysis (PFA).

|  | **factor** |  |  |  |
| --- | --- | --- | --- | --- |
| **z score variables** | **1** | **2** | **3** | **4** |
| **STAI-T<sup>a</sup> sum score** | 0.9 |  |  |  |
| **CSQ-D<sup>b</sup> catastrophizing** | 0.9 |  |  |  |
| **CES-D<sup>c</sup> sum score** | 0.8 |  |  |  |
| **PCS-D<sup>d</sup>** | 0.8 |  |  |  |
| **STAI-S<sup>e</sup> sum score** | 0.8 |  |  |  |
| **rel. gene expression IL10<sup>g</sup>** |  |  |  |  |
| **CSQ-D<sup>b</sup> self-instructions** |  | 0.8 |  |  |
| **CSQ-D<sup>b</sup> distraction** |  | 0.7 |  |  |
| **CSQ-D<sup>b</sup> ignore** |  | 0.7 |  |  |
| **CSQ-D<sup>b</sup> reinterpretation** |  | 0.6 |  |  |
| **CSQ-D<sup>b</sup> praying hoping** |  | 0.5 |  |  |
| **CSQ-D<sup>b</sup> activity increase** |  | 0.5 |  |  |
| **CSQ-D<sup>b</sup> pain control** |  | 0.4 |  |  |
| **CSQ-D<sup>b</sup> pain reduction** |  | 0.3 |  |  |
| **GCPS-D<sup>h</sup> pain intensity** |  |  | -0.7 |  |
| **GCPS-D<sup>h</sup> grade** |  |  | -0.6 |  |
| **GCPS-D<sup>h</sup> disability** |  |  | -0.6 |  |
| **NPSI-D<sup>i</sup> sum score** |  |  | -0.6 |  |
| **rel. gene expression IL4<sup>i</sup>** |  |  |  |  |
| **CSQ-D<sup>b</sup> pain behavior** |  |  |  | -0.6 |
| **rel. gene expression TNF<sup>k</sup>** |  |  |  | 0.5 |
| **rel. gene expression IL6<sup>l</sup>** |  |  |  | 0.3 |
| **$\alpha^m$** | 0.9 | 0.8 | 0.7 | 0.5 |

<sup>a</sup>STAI-T = subscale trait-anxiety of the German version of the state/trait anxiety
inventory
<sup>b</sup>CSQ-D = subscales of the German version of the coping strategies questionnaire
<sup>c</sup>CES-D = German version of the center of epidemiological studies general depression
scale
<sup>d</sup>PCS-D = German version of the pain catastrophizing scal
<sup>e</sup>STAI-S = subscale state-anxiety of the German version of the state/trait anxiety
<sup>f</sup>IL10 = interleukin 10
<sup>g</sup>GCPS-D = three subscales of the German version of the graded chronic pain
scale<sup>h</sup>NPSI-D = German version of the neuropathic pain scale inventory

The items that load on the same factors suggest that factor 1 represents “affective load”, factor 2 “coping strategies”, factor 3 “physical functioning” and factor 4 “pro-inflammatory cytokines”. The loadings are given in descending order representing that anxiety and catastrophizing are the items that have the most decisive influence on variance (Table 1). Followed by self-instructions / reinterpretation / praying and hoping, pain intensity and pain behavior.

### Affective load, coping, physical functioning and cytokine profile cluster patients into four subgroups

A hierarchical cluster analysis was performed on the individual factor scores to distinguish patient subgroups. We decided to interpret a four-cluster solution as the best fit for our cohort (see S8 Fig). Finally, we characterized and labeled these clusters based on differences regarding the factor scores as following: Cluster A as “maladaptive”, cluster B as “adaptive”, cluster C as “vulnerable” and cluster D as “resilient”. The differences in the mean factor scores at the four profiles are illustrated in Fig 2 and their scores are compared in S3 Table.

**Fig 2:**
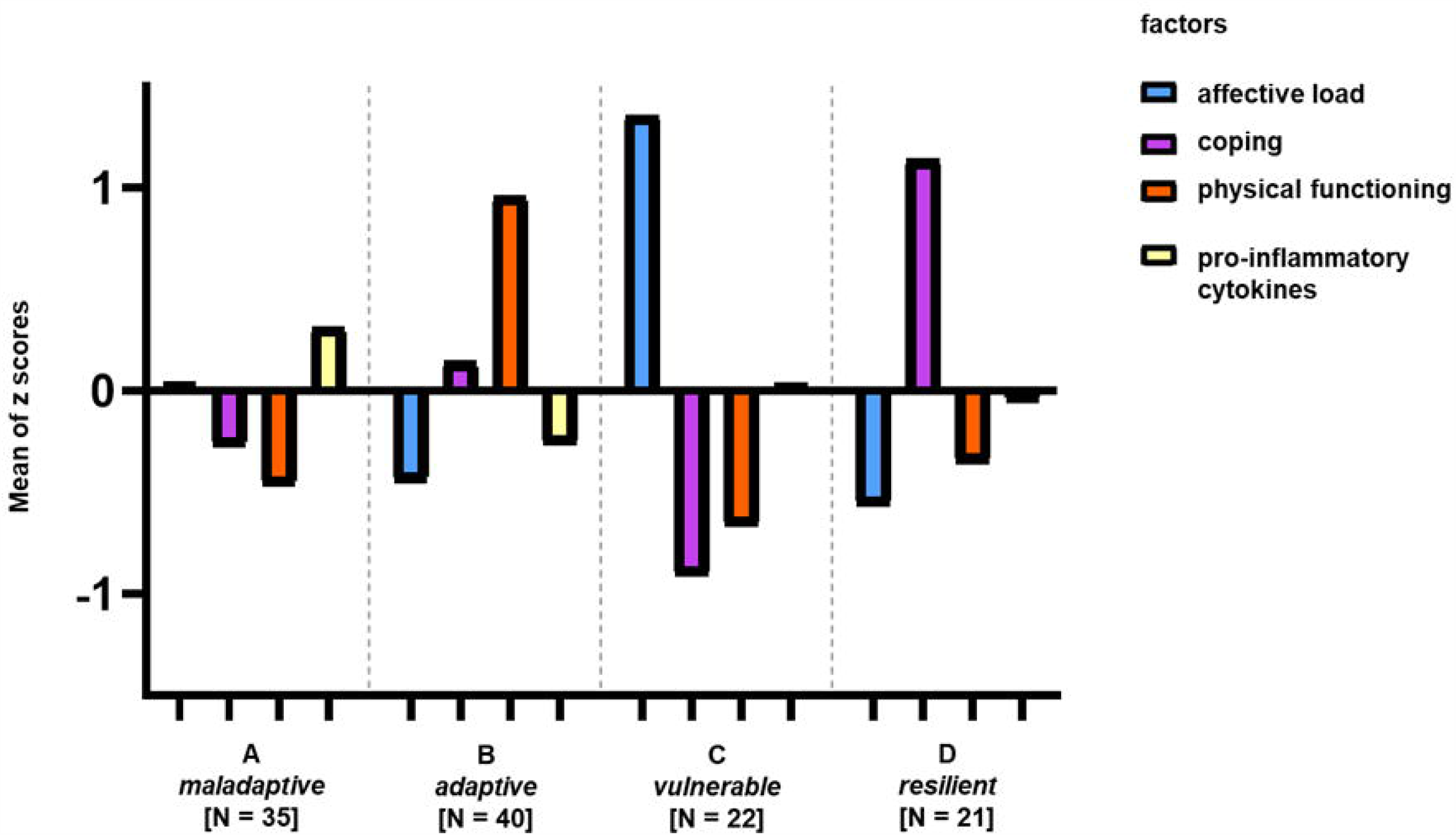
Four clusters differentiated by four factors explaining the variance in somatic and psychosocial data within the patient group. Bars symbolize positive or negative difference of this cluster in one specific factor to the mean value in this factor of the entire group. No bar symbolizes no difference of the group in this factor to the mean value of the factor of the entire group. Cluster A (named “maladaptive”) consists of 35 patients, the “adaptive” cluster of 40, the “vulnerable” cluster of 22 FMS patients and cluster (named “resilient”) of 21 patients. Significant differences between factors and groups are marked (p < 0.05). Abbreviations: FMS = Fibromyalgia Syndrome.

A 3D model provides a more convenient illustration of the four clusters characterized by four factors and is available from following link: https://drive.google.com/file/d/14r8DXQN1akmftBSYWWFRNv3sZlTQvtpI/view?usp=sharing [49].

The cluster labels are based on the differences and characteristics of each individual cluster which described in the following. The maladaptive cluster A (N = 35) is characterized by negative affect, maladaptive coping, low scores in physical functioning and a higher gene expression of pro-inflammatory cytokines (p < 0.05, compared to cluster B). Maladaptive coping might be dysfunctional and not effective to maintain physical functioning. The adaptive cluster B (N = 40) includes patients with a low negative affect who are using adaptive coping to deal with the highest pain scores (p < 0.05 compared to the vulnerable cluster C; see S3 Table), and lower values for pro-inflammatory cytokines. The active way of coping has positive effects on physical functioning. Cluster C (“vulnerable”, N = 22) is characterized by high negative affect and destructive coping (p < 0.05). Cluster D (“resilient”, N = 21) has the lowest scores in affective load (p < 0.05) and is the cluster with highest rate of active coping strategies (p < 0.05).

### Clusters differ in disease impact

To assess the impact of disease in the four different clusters, we examined differences regarding clinical disease characteristics with one-way ANOVAs (see S9 Table), followed by pairwise post-hoc comparisons (see S10 Table), and we calculated effect size values (Table 2). One-way ANOVAs revealed that clusters differed in “impact of FMS on daily activities” according to the FIQ-D sum score and in mental health according to the MCS scores (p < 0.05, Fig 3).

**Table 2:**
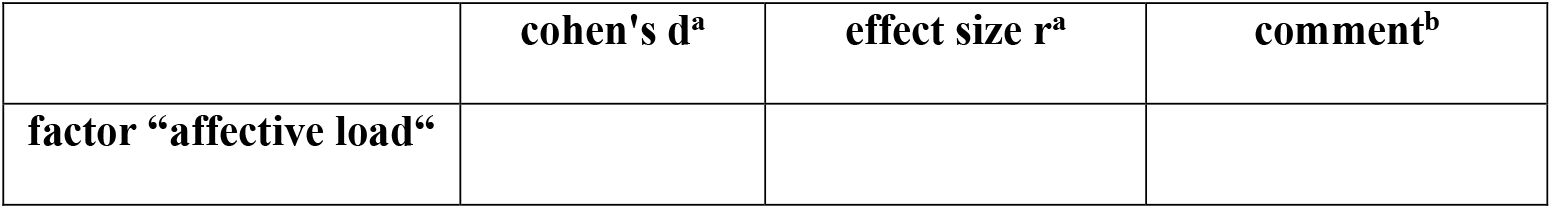
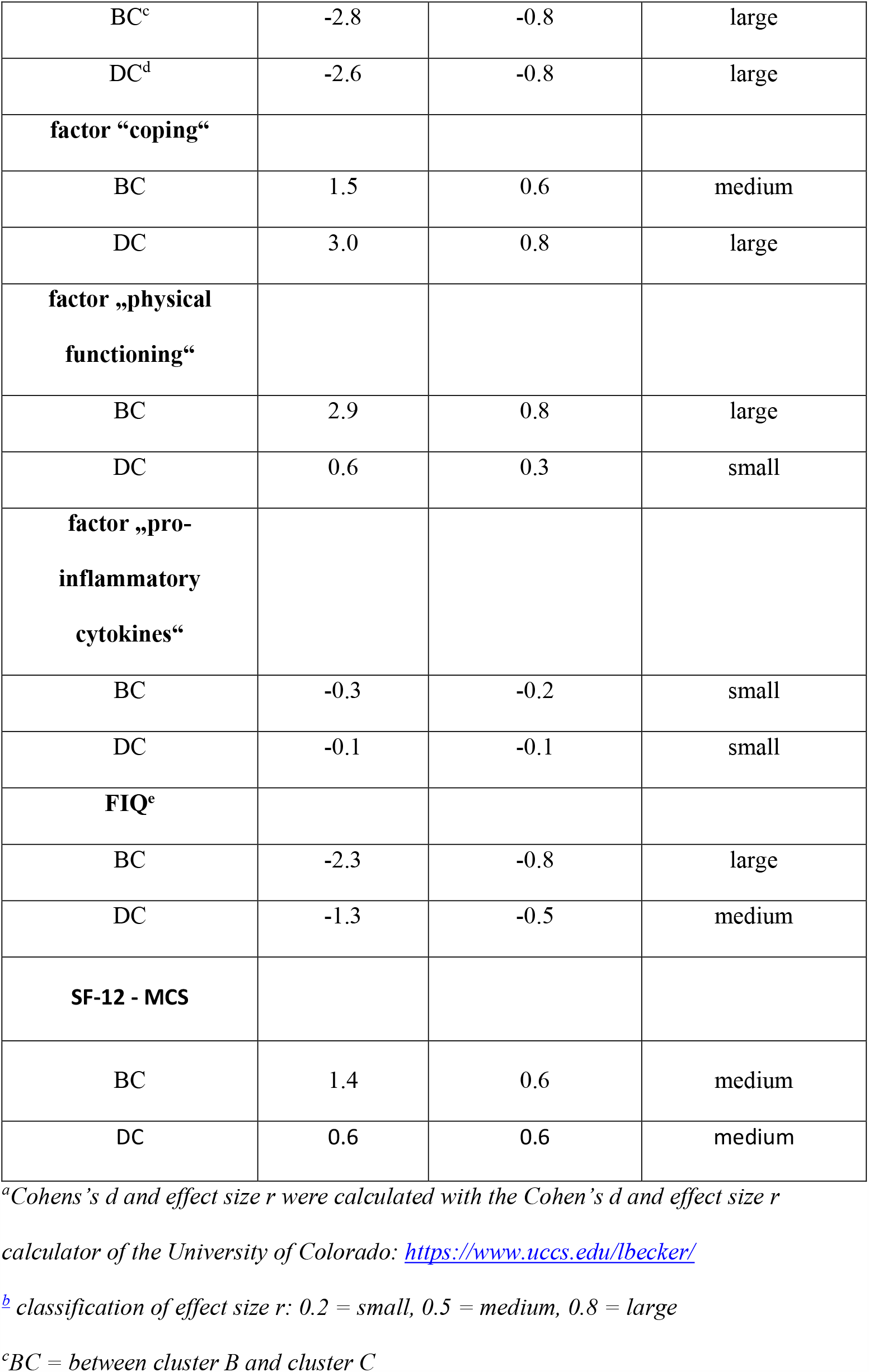
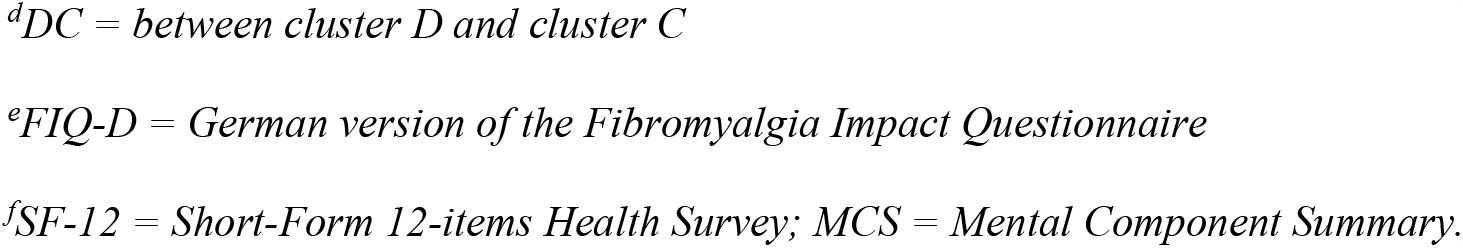
Cohen’s d and calculated effect size r between adaptive and resilient cluster B and D and the vulnerable cluster C regarding factors and some outcome variables.

**Fig 3.**
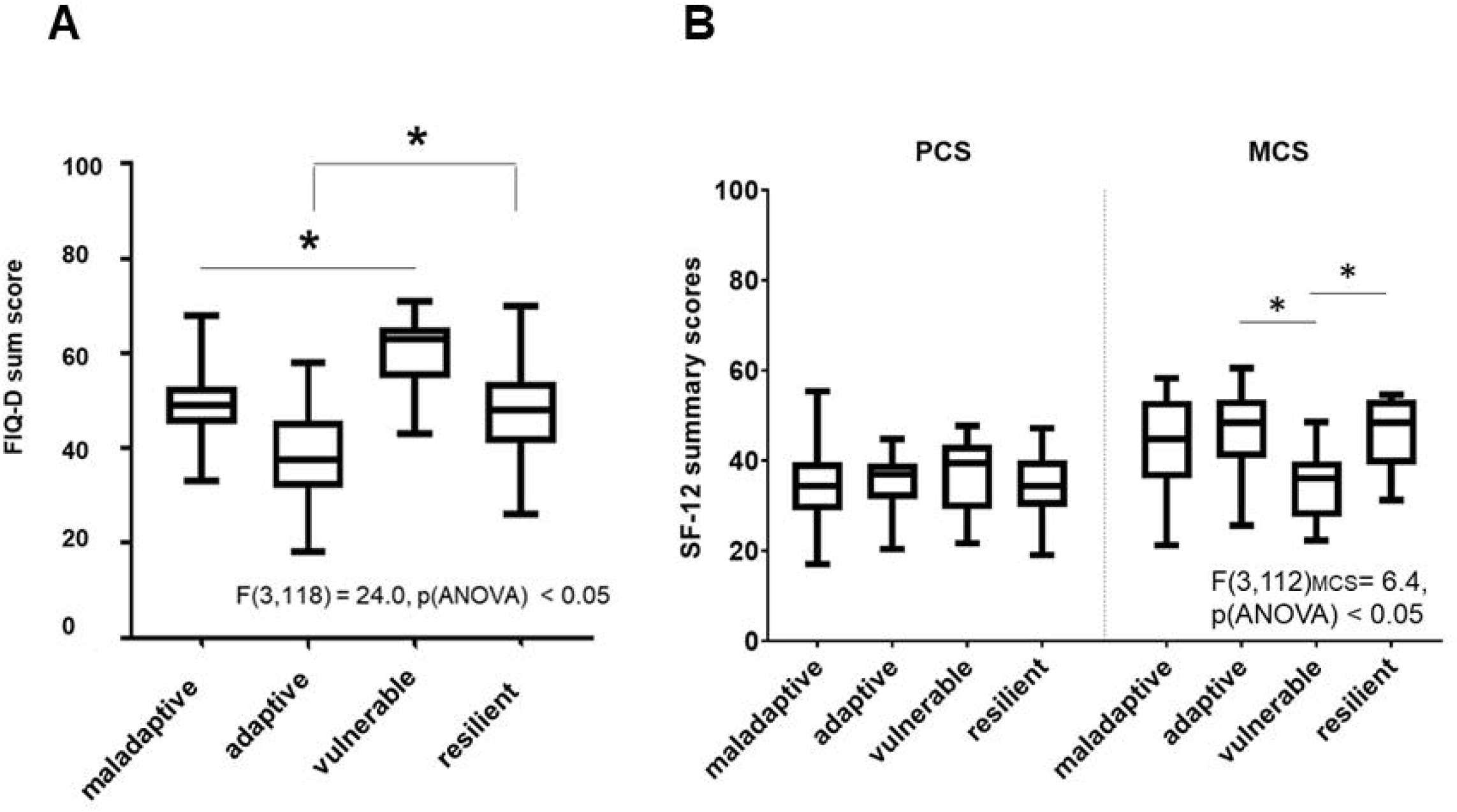
Differences of the clusters in FMS impact in life. **(A)** and self-reported quality of life (B). Boxplots of all clusters show differences in all plotted sum score variables resulting in different severity level of the clusters. Significant differences are marked (p < 0.05). Abbreviations: FIQ = Fibromyalgia Impact Questionnaire; FMS = Fibromyalgia Syndrome; SF-12 = 12-items Short-Form Health Survey; PCS = Physical Component Summary; MCS = Mental Component Summary.

The follow-up tests revealed significant differences with large effect sizes (r = −0.8; see Table 2) between cluster B vs. C, and D vs. C regarding FMS impact (p < 0.09, Fig 3A), whereas the daily life of cluster C was highly affected by FMS related symptoms and patients in this cluster had the highest disability (p < 0.05). The mental health score differed between cluster B vs. C, and D vs. C with medium effect sizes (p < 0.05), but the cluster did not differ in the physical component score.

Coping seems to be crucial for the main difference between the resilient cluster D and the vulnerable cluster C, with a large effect size between both clusters in factor 2 “coping” (r = 0.8; Table 2), whereas the effect of coping between the adaptive cluster B and the vulnerable cluster C was only medium (r = 0.6). Interestingly, the resilient cluster D had higher FIQ scores than the adaptive cluster B, suggesting that adaptive coping may be more successful than “resilient” coping. Overall, the adaptive cluster and the vulnerable cluster showed the lowest and the highest impact of disease and mental health (Fig 3, p < 0.05).

### One cluster uses reinterpretation as highly effective coping and might activate natural resources to cope with FMS

Cluster specific coping patterns, which might indicate vulnerable or resilient coping, were evaluated based on the mean using frequencies of the eight different behavioral and cognitive coping strategies and their subjective efficacy as assessed with the CSQ-D (Fig 4).

**Fig 4.**
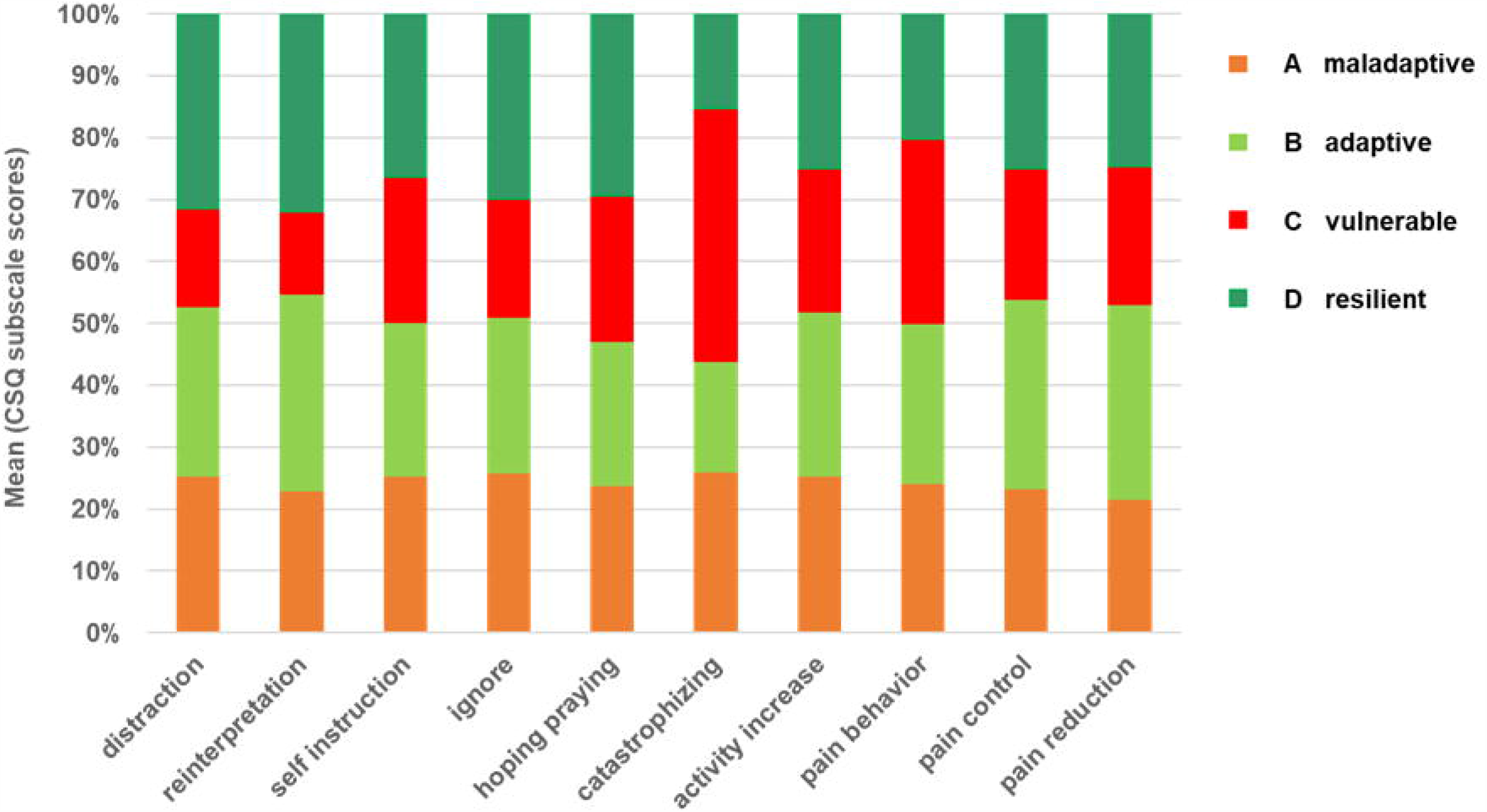
Differences between clusters regarding coping strategies. Bars illustrate the different proportion of the clusters A to D on the mean value of 8 subscale scores of the CSQ-D questionnaire indicating different coping strategies and 2 scores indicating efficiency of these strategies. The coping pattern of the vulnerable cluster is characterized by the high impact of catastrophizing (p < 0.05), the coping of the maladaptive cluster shows higher values in less effective passive strategies with relatively high values in catastrophizing, the adaptive clusters copes with effective problem and emotion-focused strategies with high values in pain control and reduction (p < 0.05) and the resilient cluster uses cognitive based coping strategies with high efficiency in pain control. Abbreviations: CSQ = Coping Strategies Questionnaire.

The group differences between all coping strategies including pain control and pain reduction were significant (p < 0.05) except for “pain behavior”. The coping type of patients within cluster C might be described as vulnerable coping, since this cluster had the highest frequency in catastrophizing (M = 25.9, SD = ±5.0) and passive negative coping strategies with less effect on pain control (MED = 2.0 (0 – 3) and symptom reduction (MED = 2.0 (0 - 4) (S3 Table). Resilient coping might be seen in cluster D, which had the highest values in all active coping strategies especially in reinterpretation with the most positive effect on pain control (MED = 4.0 (0 - 5)) and pain reduction (MED = 2.0 (0 – 4)) compared to the maladaptive and vulnerable cluster (Fig 4, Table 2, S3 Table; p < 0.05). Reinterpretation or reappraisal is a well-known cognitive technique to modulate and control negative emotions resulting in reduced subjective experience of emotions and physiological response. The ability to cope with this technique normally needs professional support. Thus, cluster D seemed to be naturally prepared to cope effectively with pain and other symptoms indicating the ability to activate resources which is also defined as resilience.

### Cluster specific cytokine pattern

Effect sizes of factor 4 (“pro-inflammatory cytokines”) between the adaptive (B) and the vulnerable cluster C (BC; r = −0.2), and between the resilient cluster (D) and the vulnerable cluster C (DC; r = −0.1) were small, suggesting that differences in gene expression of all measured cytokines regarding the cluster categories were minor (Table 2), which was confirmed by the data in Table 2. Searching for a specific cytokine pattern related to resilience, we only found differences between cluster D versus C and cluster D versus A regarding the anti-inflammatory gene target IL-10 (Fig 5; p < 0.05).

**Fig 5.**
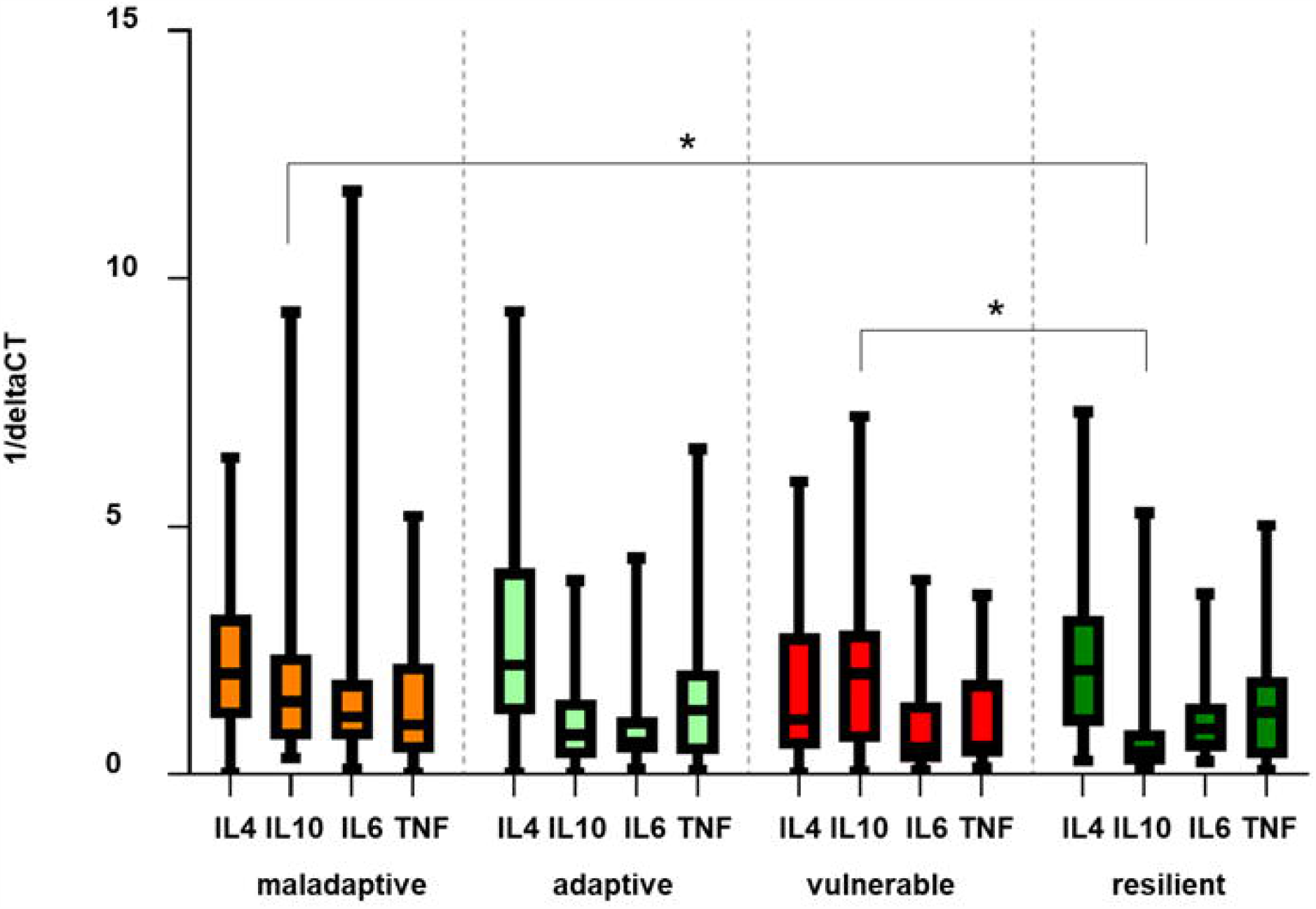
Cluster specific cytokine profiles. Bars illustrate mean deltaCT values of the target normalized to the housekeeping gene 18sRNA, of IL-4 (blue), IL-6 (red), IL-10 (green) and TNF (orange) of patients with FMS organized by the four defined clusters. Results are presented as 1/ΔCT. Relative gene expression of IL-10 between cluster A and D, and between cluster C and D are marked as significant (p < 0.05). Abbreviations: FMS = Fibromyalgia Syndrome; IL = Interleukine; TNF = Tumor necrosis factor-alpha.

All other comparisons only revealed a trend toward lower levels of pro-inflammatory cytokines in the adaptive and the resilient cluster, such that a specific cytokine pattern related to resilience could not be verified.

### Cluster – specific case description

To illustrate the findings in our cluster analysis, we selected one typical patient per cluster and described their social environment and socio-demographic data. The case descriptions underline and discuss the cluster labels (Fig 6).

**Fig 6.**
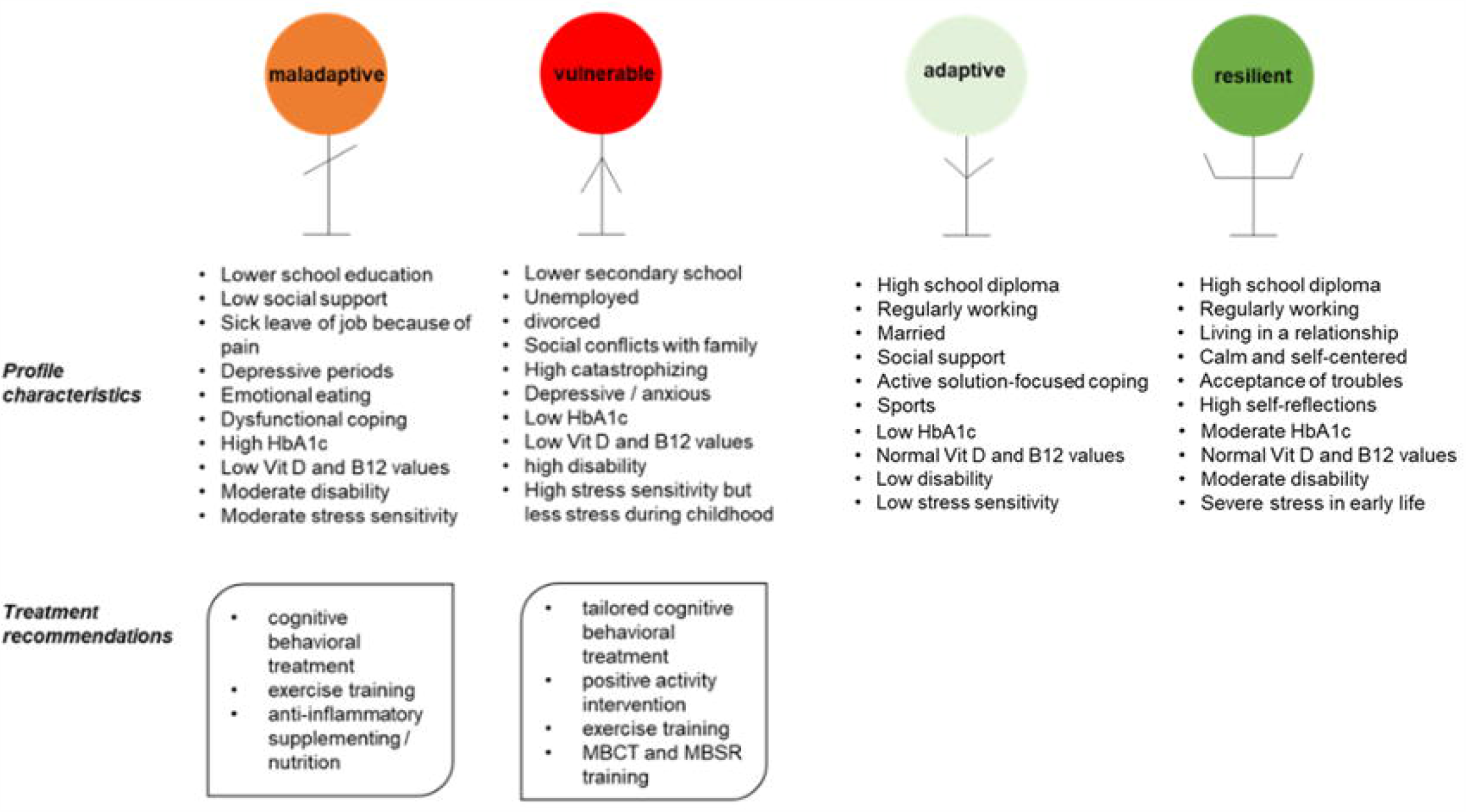
Cluster-specific case description and treatment options. Four profiles of FMS patients per cluster were selected, and possible interventions are listed based on cluster characteristics and missing resources of the vulnerable and maladaptive cluster to improve the severe and maladaptive phenotypes. Abbreviations: MBCT = Mindfulness-Based Cognitive Therapy. MBSR: Mindfulness-Based Stress Reduction. Additional comments describing the subjective personal impression on each patient were taken during the medical history and the psychological interview on the study day (see S2 Fig).

The typical “maladaptive” patient reported experiencing neck, shoulder, and lower back pain of medium intensity for several years following no specific incident. The requirements with slight to moderate degree of difficulty were overwhelming and resulting in somatic symptoms and anxiety. Pain attacks, depressive mood, and the lack of resilient handling of stress resulting in sick leave of the job. The patient was always trying to obtain treatment (such as psychotherapy) but with little success. Finally, patient “maladaptive” resolved to unhealthy and dysfunctional coping like eating and smoking.

The typical “adaptive” patient was referred to a pain clinic after experiencing pain and accompanying symptoms like irritable bowel, headache, or concentration problems since a long time. During the treatment there, various techniques to deal with the pain were learned. The adaptive patient considered himself “a positive personality” and get a lot of support from the social environment. It demands a lot of strength to daily fight against the pain, but the adaptive patient was well able to handle daily life and to stay active in life, doing sports and listens to the own needs.

The recruitment process of the typical “vulnerable” patient was difficult, because the patient was very anxious about the study tests, seemed to be lost with the symptoms and was complaining about how bad the doctors were, that no one is helping, and everything would no longer make sense. Pain intensity was reported on the highest possible scores. The typical vulnerable patient was currently in psychological therapy that was described as exhausting, since all the work had to be done by itself instead of the therapist. The lifestyle was very inactive, no sports was done and there were many conflicts with family or friends.

The typical “resilient” patient reported intense pain and high impairment, but during history taking, the story of symptoms fades into the background. Rather, the sweltering conditions and sexual abuse during childhood resulted into uncontrolled eating habits that led to obesity which were experienced as shelter against environmental challenges. Despite the adverse experiences, good grades at school were achieved and happiness had its place in life. Working as a social educator give back some of the life experience to young people and the “resilient” patient seemed to be reflective and calm.

The commonality in each of these four cases is that the individual is experiencing pain and further symptoms that have an impact on daily life. The way to cope strongly modulates the impact.

## Discussion

We determined four factors potentially contributing to the heterogeneous clinical profiles in a well-defined cohort of 118 FMS patients. Based on these factors we were able to categorize the patients into four clusters distinguished by FMS related disability, coping, psychosocial and somatic factors, which we suggest may be related to resilience or vulnerability.

Despite advances in the understanding of pain mechanisms, chronic pain patients are still treated as a homogeneous group. A substantial proportion still has no benefits from this generic approach. Several attempts have been made to categorize chronic pain patients using different instruments (e.g. Minnesota Mental Personality Inventory (MMPI) [52]. One of the methodological problems of such one-dimensional instruments is e.g. that the MMPI is a standard instrument in psychiatric instead of medical patient cohorts. In general, most cluster studies were focused on psychosocial parameters and personality traits. So far, no data are available yet on the extent to which skin innervation (nociceptor structure and function) and nerve lesions of afferent and efferent peripheral nerve fibres affect the way of coping.

Several authors have performed cluster analyses to identify subgroups among fibromyalgia patients [22-27]. Most of the previously published cluster studies in FMS patients performed clustering without previous factoring [23, 27, 28]. We carried out PFA to reduce the number of variables, to focus more on the factors that might be responsible for vulnerability or higher resistance to adversity in our cohort, and to identify valid factors that might be responsible for the variability within the cohort.

We included a wide range of psychosocial and somatic variables, because resilience is composed of a variety of small-scale aspects. Initially, more physiological data were included into the analysis, derived from QST, PREP measurements, clinical examination and history, and skin biopsy (IENFD). In the first run, only QST and PREP appeared to have an influence on within-group variability. Unexpectedly, IENFD derived from proximal and distal leg did not emerge as loadings on any factor, although they were previously shown to relate to disease severity [58]. One possible reason could be that mental parameters such as anxiety or depression have a higher impact than parameters derived from peripheral nerves on variance among our cohort. Another point could be that nerve pathology may have an influence on the severity of the disease, but the way to cope with this severity might be decisive for the functioning in everyday life. Therefore, coping and mental parameters statistically emerge as variance influencing factors. We thus did not include QST, PREP and data of skin biopsies into the factor analysis and only focused on cytokines.

Despite the small effect sizes between the clusters in factor “pro-inflammatory cytokines”, these data influenced the variability of the cohort and were important enough to exclusively load on factor 4. If indeed higher levels of IL-4 and IL-10, as in cluster B and D, had a protective influence on pain, only cluster B would benefit from this constellation, with low disability scores and low pain scores. One caveat is that we only assessed cytokine mRNA and not protein.

With a four-factor solution including the factors (1) “affective load”, (2) “coping”, (3) “physical functioning”, and (4) “pro-inflammatory cytokines”, we achieved a clear separation of the factor loadings to thematic groups in the final run of the PFA. The highest loadings on factor 1 were anxiety and pain catastrophizing that are well-known risk factors and promote vulnerability [53, 54]. Coping items were the loadings of factor 2, leading to the name “coping strategies”. Factor 3 was clearly determined by a combination of items regarding impact of pain and FMS symptoms on daily activities resulting in the label “physical functioning”. Interestingly, the active coping strategy “pain behavior” loaded on the same factor as the pro-inflammatory cytokines, supporting the connection between pain behavior and pro-inflammatory cytokines as shown in animal models [55, 56] and in pain patients [57].

The following clustering divided the cohort into four clusters. A four-cluster solution provided the best fit for our data as in most other studies [4, 6, 7, 22, 28, 58, 59]. The al-Ándalus project [24] classified 486 FMS patients based on eight factors into five clusters named by the grade of performing as adapted, fit, poor performer, positive and maladapted. Our analysis resulted in a maladaptive, adaptive, vulnerable and resilient cluster, distinguished by the factors affective load, coping, physical functioning and pro-inflammatory cytokines. Most studies used symptom severity and variability of psychological factors to separate groups differing in resilience [25, 60], whereas in our study affective load and its control by specific coping and the cytokine balance determined the difference between resilient and vulnerable clusters. Coping is a variable that might be changed by a specific therapy and has a huge influence on physiological and psychological variables, in turn [61, 62]. The factors affective load and coping were the most discriminative factors in our cohort and decisive for a low or high FMS related disability and quality of life, confirmed by moderate to large effect sizes, indicating a minor role of somatic factors.

The choice of the right type of coping is the key to increase quality of life and resilience in aversive life periods [63, 64]. Problem-focused coping like increasing activity or seeking of social support and emotion-focused coping (e.g. positive reinterpretation) are two types of coping resulting in higher quality of life. Cluster B combines these two effective strategies (activity increase, self-instructions, ignore) resulting in low disability and ultimately in adaptation on FMS symptoms. High scores in negative affect including anxiety, depression and pain catastrophizing and the missing control of these negative emotions by coping strategies that are focused on emotions, behavioral or mental disengagement [65] suggest vulnerability in cluster C. The reason for combining specific coping strategies also depend on peoples’ experiences in childhood, on personality, inflammatory state, gender and age [63, 66-69].

Traumatic events during childhood are a well-known risk factor for FMS that also has the potential to alter brain activation patterns [70]. Cognitive coping mechanisms including reappraisal were identified to contribute to resilience in children with a history of maltreatment. Children who were able to recruit prefrontal control regions and to modulate amygdala reactivity during reappraisal had a lower risk for depression [71]. Patients in cluster D showed the highest scores in almost all types of psychoemotional stress the CTQ-D is evaluating and exclusively most frequently used reinterpretation as coping strategy. Reinterpretation or reappraisal is a well-known cognitive technique to modulate and control negative emotions resulting in reduced subjective experience of emotions and physiological response [72]. Positive emotions and cognitive reappraisal promote adaptive coping strategies and resilience [73]. The ability to cope with this technique normally needs professional support. Most patients within cluster D (8 of 21) and cluster B (18 of 40) never had an intervention by a psychotherapist or psychiatrist (see S3 Table). Thus, the subjects in cluster D who were exposed to stressful events in early life had resources to activate resilient coping strategies, therefore this cluster was termed as able to cope in a “resilient” way with the experienced adversity.

Resilience is a complex phenomenon. Many scientists regard resilience as a personality trait [74], whereas others consider it as a process of adaptation during a critical time [75, 76]. We speculate that cluster D has personal resources to actively deal with intense pain and high traumatic events, indicating a resilient trait, whereas cluster B might be best adapted and has learned how to effectively cope with FMS. The chosen labels might be debatable, but in our judgement, they reflect the different emerged profile groups in the best way. In contrast to [24], we did not include a predefined factor “resilience” by factor loadings of questionnaires such as emotional repair (TMMS-24), positive affect (PANAS) and optimism (LOT-R), focusing on the psychological ability of resistance. Our strategy was to define resilient strategies of coping with FMS that might be used to change a “non-resilient” into a more resilient phenotype and open new potential ways of treatment.

Clusters may offer the possibility for individualized therapies. Based on profile specific characteristics of both unfavorable clusters we suggest some treatment options to change a severe phenotype. These options are based on known effects on immune system and mental health published since now. Besides drug therapy there are potential alternative anti-inflammatory and microbiome-influencing therapies [12, 54, 63, 77-81] to support the immune system and mental health, cognitive behavioral and mindfulness based (MBSR / MBCT) therapies to support adaptive coping [82, 83], and exercise training with beneficial effects on fatigue, pain and mood [84, 85]. Positive activity interventions (PAI) that promote positive affect, acceptance, feeling of support and coherence and show a long-term persistent effect on pain relief and FMS related symptoms like depressive mood might be helpful [86].

General limitations of the study are that although the patient recruitment was done following strict criteria, nevertheless every study depends on suitable study participants who are willing to appear on the study day and participate on every test. Our cohort might exclude the very vulnerable individuals who were not willing or who were not able to participate on a series of tests on a specific day in the clinic far away from their familiar environment.

## Conclusions

Our FMS patient cohort consists of four clusters defined as maladaptive, adaptive, vulnerable, and resilient. Crucial for the resilient cluster were resilient coping with focus on cognitive strategies like reappraisal, associated with low depression scores, low values in other psychopathological symptoms despite the high frequent experience of high traumatic stress in early life, and high relative gene expression of the anti-inflammatory cytokine IL-10 compared to the vulnerable cluster. Resilient coping seems to consist of a combination of personal characteristics and learned behavior and might be trained based on the described coping pattern of the resilient cluster.

## Data Availability

All data relevant for this manuscript are available in the submitted manuscript.

## Acknowledgments

Expert statistical support by Dr. T. Raettig, expert technical help by D. Urlaub, and expert illustration of the 3D model by Dr. Jérémy Signoret-Genest is gratefully acknowledged.

## Supporting information

**S1 Table. Exclusion and inclusion criteria of patient recruitment**.

**S2 Fig. Overview of the measurements during the larger study on FMS**.

**S3 Table. Sociodemographic, electrophysiological, laboratory, psychosocial, and somatic characteristic differences among cluster**.

**S4 Fig. Flow chart of patient recruitment of this study**.

**S5 Table. Adequacy tests of the principal axis factoring analysis**.

**S6 Table. Data of variance of emerged factors with eigenvalues more than 1 observed between predicting variables**.

**S7 Fig. Scree plot (A) before and (B) after predefining the number of factors. S8 Fig. Dendrogram of the cluster analysis and the marked four cluster**.

**S9 Table. One-way ANOVA to test the significance between factors between the subgroups**.

**S10 Table. Post-hoc analysis between subgroups and factors**.

